# Mortality Classification for Deaths that Follow the Use of Non-Firearm Force by Police: A National Cross-Sectional Study (United States, 2012-2021)

**DOI:** 10.1101/2024.05.20.24307634

**Authors:** Justin M. Feldman, Tracey Lloyd, Phillip Atiba Solomon

**Affiliations:** Center for Policing Equity; François-Xavier Bagnoud Center for Health and Human Rights, Harvard University; Department of African American Studies, Yale University; Department of Psychology, Yale University

## Abstract

**Background:** Mortality classification for deaths in US police custody has important consequences for epidemiologic monitoring and legal outcomes. Prior literature suggests in-custody death classification is inconsistent and may not reflect non-firearm force that preceded death.

**Methods:** We analyzed the Associated Press “Lethal Restraint” national dataset (United States, 2012-2021; N = 1,036), which included deaths following police use of non-firearm force. Our primary outcomes included whether the death investigator: (1) classified manner of death as a homicide, (2) mentioned a force-related injury/condition in the cause-of-death statement, and (3) mentioned any force. Inverse-probability-weighted logistic models estimated the association of these outcomes with death-investigator jurisdiction type, local political composition (quartile of Republican Party vote %), decedent race/ethnicity, and each agency’s prior classifications.

**Findings:** We removed 96 deaths based on exclusion criteria. Of the remaining 940 deaths, 28.5% were classified as homicide, 16.5% had cause-of-death statements mentioning a force-related injury/condition, and 42.6% mentioned any force. In contrast, 73.9% of statements mentioned drugs. Unadjusted results showed homicide classification increased from 25.0% in 2012-2014 to 32.2% in 2018-2021. Models estimating adjusted prevalence differences (aPD) showed that, compared to medical examiner jurisdictions, coroners (aPD: -0.19; 95% CI: -0.31, - 0.06) and sheriff-coroners (a PD: -0.17; 95% CI: -0.28, -0.05) were less likely to classify deaths as homicides. Model results also showed that classifications for incidents occurring in the least-Republican counties were most likely to reflect force across all three manner and cause outcomes.

**Interpretation:** Non-homicide classifications and cause-of-death statements making no mention of force were widespread for US in-custody deaths. We identified novel evidence suggesting coroner and sheriff-coroner jurisdictions were especially unlikely to categorize in-custody deaths as homicides, and that incidents occurring in highly Republican counties were least likely to reflect force in the cause or manner of death.

**Funding:** Blue Meridian Partners Justice and Mobility Fund

## Introduction

During the past decade, deaths occurring after police use-of-force have garnered considerable attention. Subsequent public concern about police accountability has extended to public health and medicine: Statements by US professional associations in these fields have called for improved investigation and data reporting for police-related deaths,^1,2^ which are often misclassified in official mortality data.^3^ Deaths following the use of non-firearm force by police (e.g., chokeholds, prone restraint, or conducted energy weapons (CEWs)) are particularly likely to be classified inconsistently; classifications of these deaths may fail to reflect the violent confrontation that preceded the death. ^3–5^ Mortality classifications have potential ramifications both at the level of an individual death (e.g., for determining whether a police officer caused a death in legal proceedings) and systemically (e.g., for monitoring social inequities in death rates or assessing the safety of police restraint practices).^6^ Despite the salience of this issue, investigations into the classification of non-firearm force-related deaths have been primarily limited to journalistic accounts, opinion surveys of forensic pathologists, and statistical analyses of small convenience samples of mortality data.^3–5,7^

In the US, medical examiners (appointed officials who are typically forensic pathologists) and coroners (generally, elected officials without medical training) investigate deaths in custody and make official determinations about the manner and cause of death. Manner of death is a categorization of the circumstances under which a death occurred, and includes natural, accidental, suicide, homicide, and undetermined (Table 1). While a homicide determination does not imply that an unlawful act occurred, it does indicate that law enforcement actions played a causal role in the death, either through use-of-force, medical neglect, or engaging the subject in a physical struggle that led to death via pathways of stress and exertion. The National Association of Medical Examiners, the primary US professional association for forensic pathologists, suggests classifying deaths resulting from police subdual or restraint as homicide,^1^ but prior evidence indicates death investigators routinely select non-homicide manners of death in these scenarios. In contrast, a cause of death is a written statement of the diseases, injuries, or complications that directly caused or contributed to a death. Cause-of-death statements for high-profile incidents have included force-related injuries or conditions (e.g., the medical examiner’s mention of “neck compression” in the cause of death statement for the 25 May, 2020 death of George Floyd),^8^ but these cases may be exceptional. Prior evidence suggests that a potentially large proportion of deaths following non-firearm force exclusively identify cardiovascular conditions, drugs, and/or psychiatric illness as causes.^3^

**Table 1.**
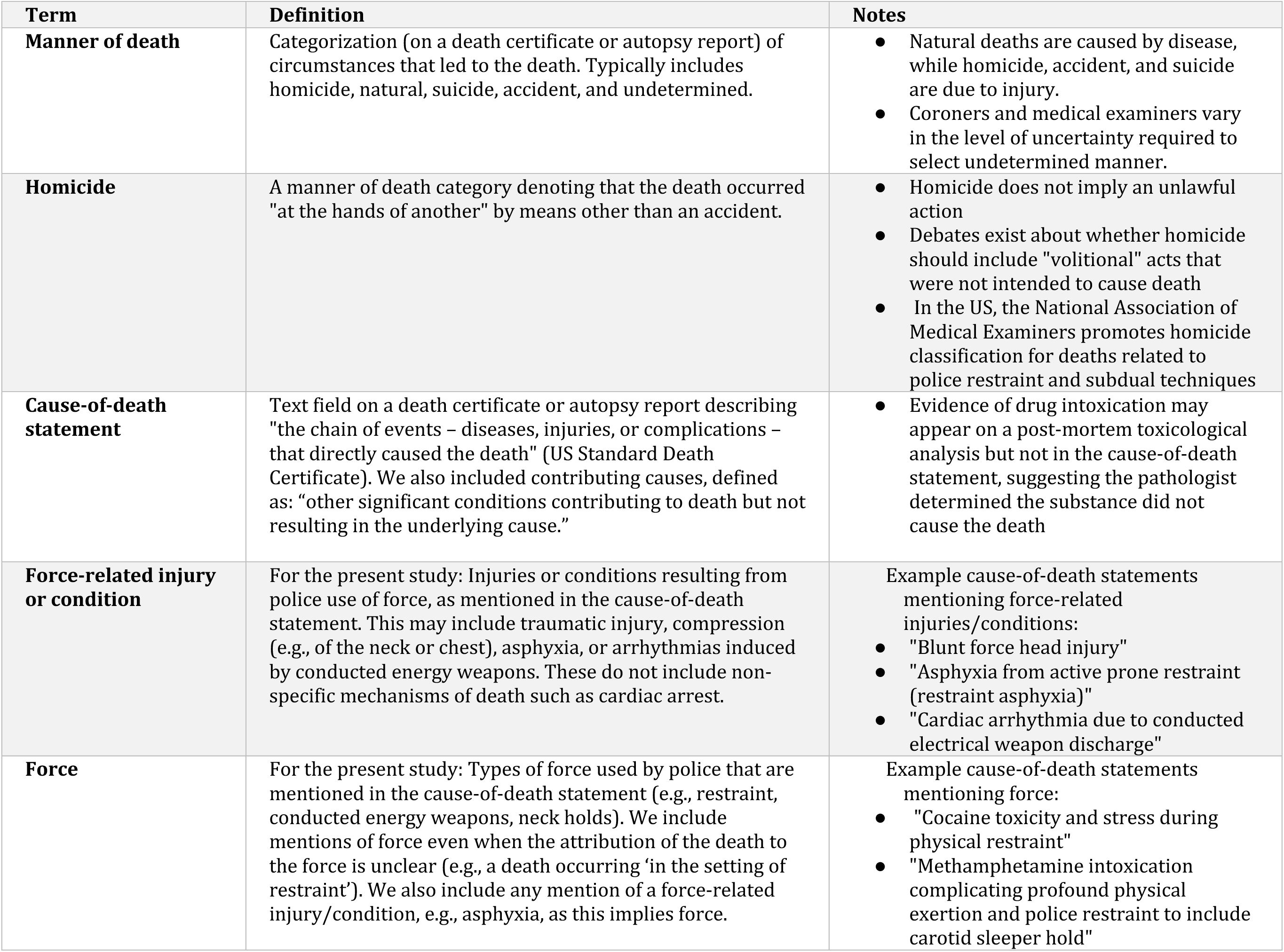
Description of death categorization terms.

Various factors may lead investigators to classify deaths in ways that fail to reflect the involvement of police use-of-force. First, causation for deaths in police custody is often multifactorial: Deaths often involve drug intoxication, physical exertion, and underlying chronic disease.^1^ The consequences of force applied in these settings are often unclear, with autopsies and post-mortem toxicological analysis typically failing to yield definitive conclusions.^9^ Further complicating the determination of deaths in police custody, some forensic pathologists cite a contested condition called “excited delirium” or “agitated delirium” – characterized by hyperactive, agitated behavior and often related to drug intoxication – as a cause of deaths in police custody, and may believe that the role of restraint maneuvers in such deaths is incidental.^10^ Second, idiosyncratic differences, reflecting variation in pathologists’ training and preferences, affect death classification practices.^5^ For example, there are varying schools of thought about whether assigning homicide as a manner of death requires intent to kill.^5,11^ Third, conflicts of interest may play a role both in the death investigation process itself and in the research literature that informs forensic pathologists’ conclusions about deaths that follow non-firearm force. In some jurisdictions, a sheriff or other law enforcement official serves as the coroner; these individuals may be charged with investigating their own agency’s actions.^12^ Additionally, researchers and journalists have described conflicts of interest in the scientific literature assessing the safety of police restraint techniques and technologies, as the authors of such literature may also serve as expert witnesses, perform liability-avoidance trainings for police administrators, and receive research funding from police technology companies.^13,14^ Furthermore, prior research suggests that anti-Black racial bias may affect the classification of injuries and injury-related deaths, including those occurring in jails, although to our knowledge, no research has examined racial bias for the classification of deaths in police custody.^4,15^ Finally, local partisan politics may play a role in death determinations. Prior research suggests that COVID-19 deaths were more likely to be misclassified in areas with stronger support for the Republican Party.^16^ In-custody deaths are also highly politicized, and classification may reflect higher perceptions of police legitimacy reported among US Republicans versus Democrats.^17^

In the present study we provide what is, to our knowledge, the first national analysis of cause and manner classification for non-firearm deaths in US police custody. To do so, we leverage the Associated Press (AP News) Lethal Restraint database: national dataset, published in April 2024, containing details about deaths that follow the use of non-firearm force by police in the United States during the period 2012-2021.^18^ Our study aims to quantify the proportion of these fatal incidents whose cause and manner of death classifications reflect the involvement of police use-of-force. We also test four hypotheses — informed by prior literature on the roles of death-investigator type, political partisanship, anti-Black racial bias, and idiosyncratic variation in mortality reporting — on determinants of classification decisions for in-custody deaths. We hypothesize that cause and manner classifications will more likely reflect use of force in medical examiner jurisdictions (vs. coroner and sheriff-coroner jurisdictions), in counties with low Republican Party voting concentrations (vs. those with high concentrations), for non-Hispanic white decedents (vs. non-Hispanic Black decedents), and for death investigation agencies that have previously indicated force-involvement in their mortality classifications of in-custody deaths (vs. those that have not indicated force involvement for previous deaths).

## Methods

Our study analyzed the Lethal Restraint dataset published by AP News in April 2024.^18^ The dataset contains details of 1,036 deaths that followed the use of non-firearm force by state and local law enforcement officers including, according to AP News, approximately 270 deaths that had not been reported in previous databases.^19^ We added to this dataset by classifying the jurisdiction type (i.e., medical examiner, coroner, sheriff-coroner) for each death and added county-level variables based on the incident location. In addition to descriptive tabulations, we estimated inverse propensity-weighted logistic regression models to assess our hypotheses, which adjusted for a set of potential confounders related to decedent demographics, force types used, and geographic context.

### Data Sources

Our principal data source for the study was AP Lethal Restraint, whose methods are described in greater detail on the source website.^19^ In brief, AP News journalists conducted an open-source search of deaths that followed use of non-firearm force by state or local police that occurred in the 50 US states and District of Columbia during the period 2012-2021. They excluded deaths occurring in jails or prisons, deaths solely involving federal law enforcement, and incidents in which handcuffs were the only type of force applied. The authors did not make judgements as to whether force caused a death; instead, they included deaths that were temporally associated with force, which typically included those who died during the police encounter or during a hospital stay that immediately followed the incident. The resulting dataset includes, inter alia, decedent demographics, force types, cause of death, and manner of death as well as the names of the death investigation offices and law enforcement agencies involved. Although the Lethal Force data are publicly available on the AP News website, the journalists made additional variables (contributing causes of death; source document types for cause and manner) available to us for the current study.

AP News journalists obtained data for Lethal Restraint from official documents, personal communications with public officials, and – for force types – reviews of video footage when available. Cause and manner of death were most often obtained from autopsy reports (accounting for the source in approximately 60% of deaths), but also included personal communications with death investigators and documents from state police investigations that reference pathologist determinations. Official death certificates were only available as a source document for <10% of deaths due to legal barriers to death certificate access.

Using the Lethal Restraint data, we geocoded incident locations (reported as cities by AP News) to counties, then added county poverty rate data from the US Census American Community Survey, 5-year estimates as well as urbanicity categories using the US Department of Agriculture’s 2023 Rural-Urban Continuum Codes.^20^

### Outcomes

To characterize whether a death’s classification reflected police use-of-force, we created three binary outcome categories based on the cause-of-death statement and manner of death (Table 1):

1. Homicide: The death was classified as a homicide.
2. Mention of force-related injuries or conditions: The cause-of-death statement mentioned specific force-related injuries or conditions, including traumatic injury (e.g., fractures, wounds, blunt force trauma) as well as asphyxia, compression (e.g., of the neck or chest), or cardiac conditions related to CEWs. We categorized the latter as a force-related condition only when the weapon was also mentioned, e.g., ‘CEW-induced arrhythmias’, as the same cardiac conditions may result from other chains of events, such as drug overdose.
3. Any mention of force: The cause-of-death statement made any mention of force, e.g., restraint, CEWs, tackle maneuvers, or neck holds. For this category, we interpreted force-related injuries/conditions as implying use-of-force so, e.g., asphyxia would be considered a mention of force even if the restraint that caused the asphyxia was unmentioned. As a result, all deaths in category 2 above are included here.

We additionally categorized cause-of-death statements in terms of whether they mentioned drugs and whether they mentioned excited or agitated delirium. For all cause of death categories, two research analysts reviewed the cause-of-death statements and reached consensus for cases in which categorization was initially discordant. All cause-of-death categorizations were also based on any contributing causes reported by the death investigator.

### Exposures

We classified death-investigator type (medical examiner, coroner, or sheriff-coroner) for each death based on the type of office that had legal jurisdiction over the death. In the sheriff-coroner category, we also included a small number of agencies that were substantially similar to sheriff-coroners, including jurisdictions for which a police chief served as coroner (n = 4 deaths) and a coroner that relied on a forensics agency operated by the local sheriff (n = 5 deaths).

Although the name of the agency that performed the autopsy was available in the AP Lethal Restraint data, in many cases this was not the same agency with legal jurisdiction over the death investigation due to local outsourcing arrangements. We therefore inferred legal jurisdiction and classified office types by following a set of decision rules, which included relevant statutes in the state of death, the names of other death investigation offices identified in the AP Lethal Restraint dataset, death investigation office websites, the location of the county in which the use-of-force incident occurred, and personal communications with death investigation offices. We present the classification methods in greater detail in the supplementary material.

We measured the concentration of Republican Party voters based on the percent of bipartisan votes that were for the Republican presidential candidate in the incident county (i.e., Republican vote count / (Republican + Democratic vote count)). We calculated this variable using county-level data from the MIT Election Lab for 2012, 2016, and 2020.^21^ Each death received a partisanship value based on the presidential election that occurred in the year of death or, if not in a relevant election year, during the most recent presidential election.

We used the Lethal Restraint’s race/ethnicity classification to assess racial inequality in mortality classifications. AP News journalists derived race and ethnicity data from death certificates, autopsy reports, police reports, and other official documents. We combined race and ethnicity to produce joint categories, including non-Hispanic Black, non-Hispanic white, and Hispanic/Latine (of any race). We further combined all other non-Hispanic persons of color into a single race/ethnicity category due to their small sample sizes, which precluded meaningful analysis of separate categories.

To assess office-level idiosyncratic variation in death classifications, we restricted our analysis to autopsy-conducting agencies that performed at least two death investigations in the analytic dataset (N = 698 deaths that were at least the second to be investigated by the autopsy agency). If a death was autopsied by an agency that had classified at least one prior death as a homicide, we assigned the current death a 1. If the current death occurred in an agency that classified all previous deaths as non-homicide, we assigned it a 0. We then estimated the association between previous homicide classification and the current death’s classification. We repeated this process for the other two outcomes based on mention of a force-related injury/condition and any mention of force.

### Statistical Analysis

We fit separate logistic regression models for each of the three outcomes and four hypotheses, for a total of 12 models. All models were complete case analyses that adjusted for decedent age, reported force types, incident state, urbanicity of the incident county, poverty rate of the incident county, year of death, and month of death. All models also included percent Republican, jurisdiction type, and decedent race/ethnicity, but only the final three models included prior autopsy agency classifications. To relax parametric assumptions, we weighted outcome models using inverse propensity scores (estimated with “WeightIt” package in R), and continuous variables were fit using restricted cubic splines.^22,23^ Propensity scores were estimated as inverse-probability of exposure weights (method: Bayesian additive regression trees; estimand: average treatment effect on the treated). After fitting the weighted logistic regression models with standard errors clustered by autopsy agency, we used the “marginaleffects” package in R^24^ to estimate adjusted prevalence differences for each exposure-outcome combination.

## Results

In our analysis of the Lethal Restraint dataset, comprising 1,036 deaths, we excluded 11 deaths due to specific criteria: 8 involving self-inflicted wounds, 1 involving wounds inflicted by another civilian, and 2 for which gunshot wounds of the limbs were listed as contributing causes of death. We removed a further 85 deaths due to missing cause and/or manner data, yielding an analytic dataset of 940 deaths. Of these 940 deaths, 28.5% were classified as homicide, 16.5% mentioned a force-related injury/condition, and 42.6% mentioned any force in the cause-of-death statement (Table 2).

**Table 2.**
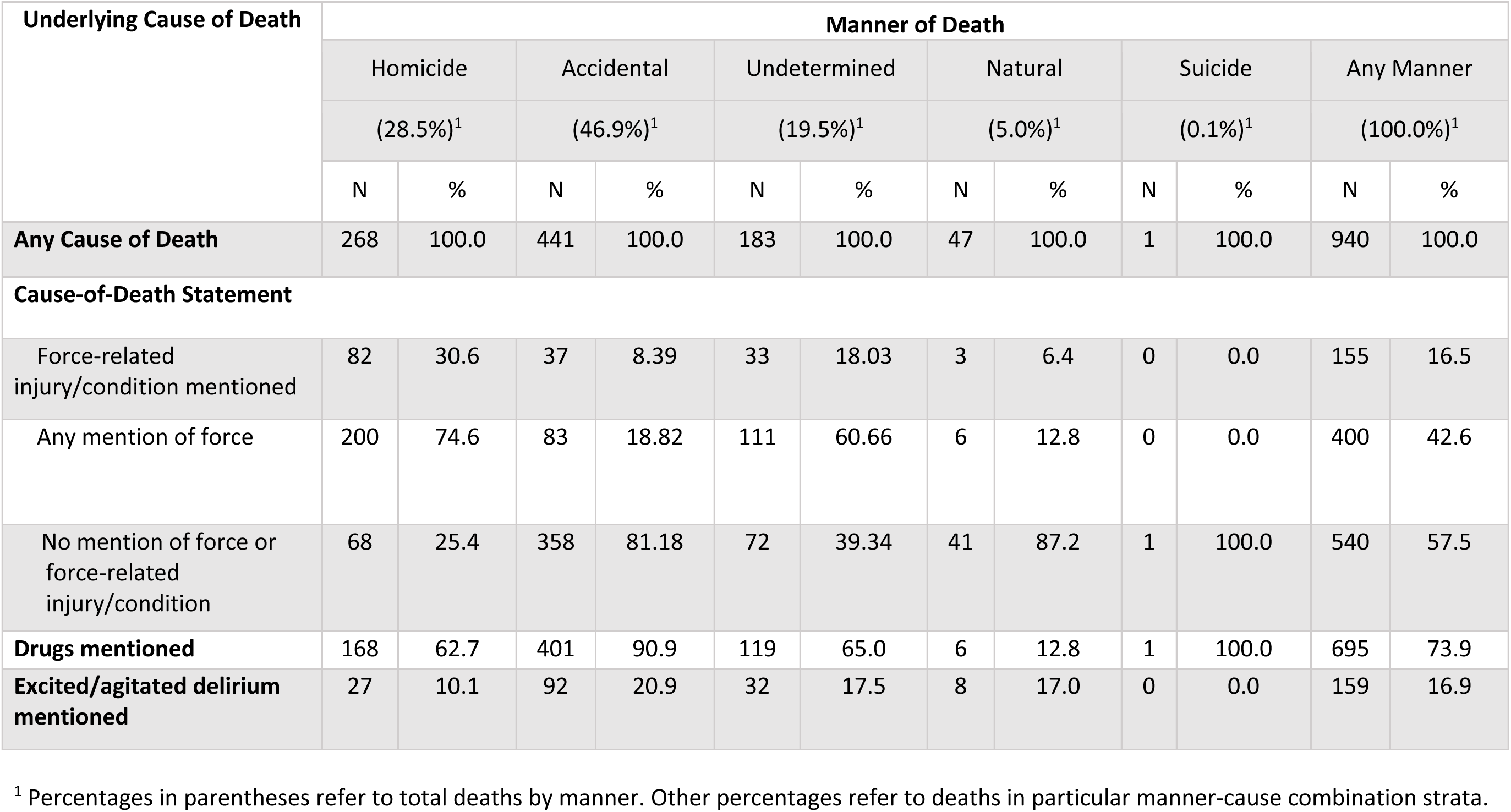
Cause-of-death statement categorizations by manner of death (N = 940 Deaths)

**Table 3.**
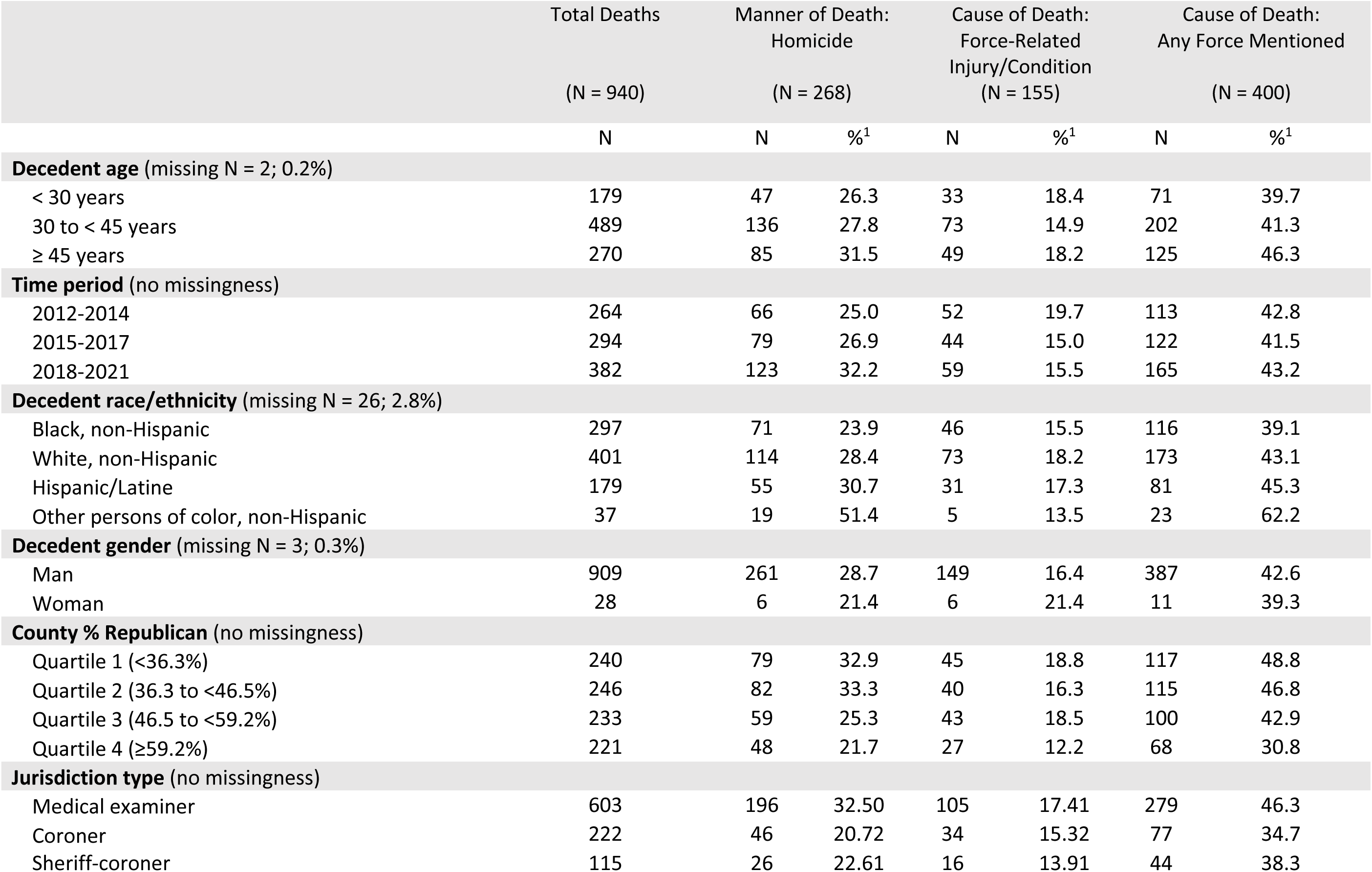
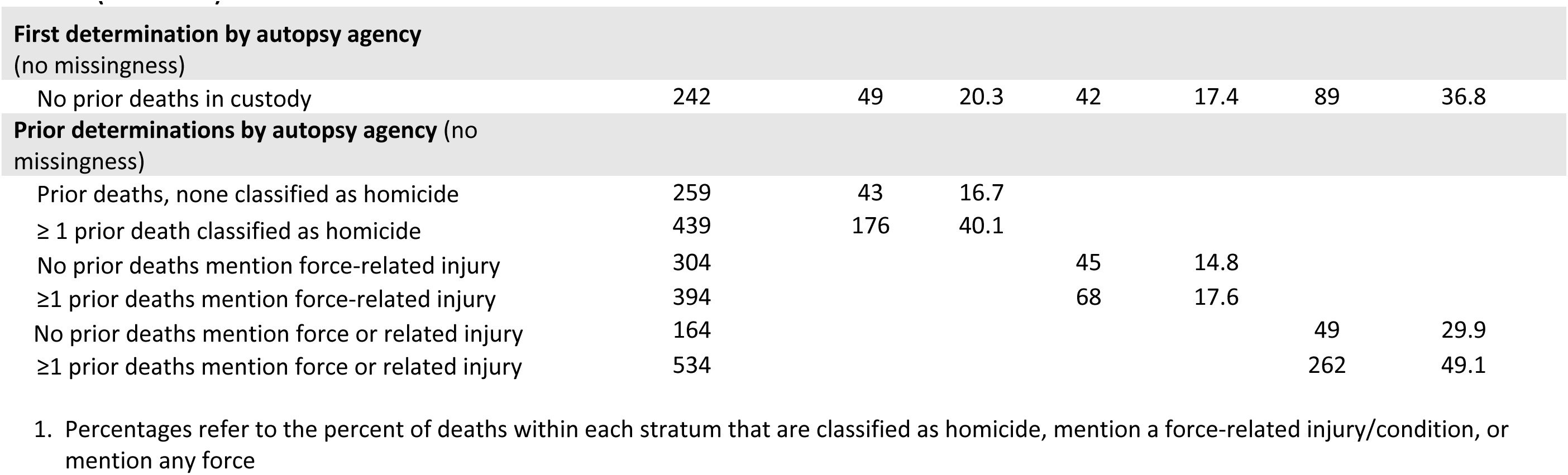
Manner and cause of death statement categorizations stratified by selected variables (N = 940 Deaths)

|  | Total Deaths<br>(N = 940) | Manner of Death:<br>Homicide<br>(N = 268) |  | Cause of Death:<br>Force-Related<br>Injury/Condition<br>(N = 155) |  | Cause of Death:<br>Any Force Mentioned<br>(N = 400) |  |
| --- | --- | --- | --- | --- | --- | --- | --- |
|  | N | N | %<sup>1</sup> | N | %<sup>1</sup> | N | %<sup>1</sup> |
| **Decedent age** (missing N = 2; 0.2%) |  |  |  |  |  |  |  |
| < 30 years | 179 | 47 | 26.3 | 33 | 18.4 | 71 | 39.7 |
| 30 to < 45 years | 489 | 136 | 27.8 | 73 | 14.9 | 202 | 41.3 |
| ≥ 45 years | 270 | 85 | 31.5 | 49 | 18.2 | 125 | 46.3 |
| **Time period** (no missingness) |  |  |  |  |  |  |  |
| 2012-2014 | 264 | 66 | 25.0 | 52 | 19.7 | 113 | 42.8 |
| 2015-2017 | 294 | 79 | 26.9 | 44 | 15.0 | 122 | 41.5 |
| 2018-2021 | 382 | 123 | 32.2 | 59 | 15.5 | 165 | 43.2 |
| **Decedent race/ethnicity** (missing N = 26; 2.8%) |  |  |  |  |  |  |  |
| Black, non-Hispanic | 297 | 71 | 23.9 | 46 | 15.5 | 116 | 39.1 |
| White, non-Hispanic | 401 | 114 | 28.4 | 73 | 18.2 | 173 | 43.1 |
| Hispanic/Latine | 179 | 55 | 30.7 | 31 | 17.3 | 81 | 45.3 |
| Other persons of color, non-Hispanic | 37 | 19 | 51.4 | 5 | 13.5 | 23 | 62.2 |
| **Decedent gender** (missing N = 3; 0.3%) |  |  |  |  |  |  |  |
| Man | 909 | 261 | 28.7 | 149 | 16.4 | 387 | 42.6 |
| Woman | 28 | 6 | 21.4 | 6 | 21.4 | 11 | 39.3 |
| **County % Republican** (no missingness) |  |  |  |  |  |  |  |
| Quartile 1 (<36.3%) | 240 | 79 | 32.9 | 45 | 18.8 | 117 | 48.8 |
| Quartile 2 (36.3 to <46.5%) | 246 | 82 | 33.3 | 40 | 16.3 | 115 | 46.8 |
| Quartile 3 (46.5 to <59.2%) | 233 | 59 | 25.3 | 43 | 18.5 | 100 | 42.9 |
| Quartile 4 (≥59.2%) | 221 | 48 | 21.7 | 27 | 12.2 | 68 | 30.8 |
| **Jurisdiction type** (no missingness) |  |  |  |  |  |  |  |
| Medical examiner | 603 | 196 | 32.50 | 105 | 17.41 | 279 | 46.3 |
| Coroner | 222 | 46 | 20.72 | 34 | 15.32 | 77 | 34.7 |
| Sheriff-coroner | 115 | 26 | 22.61 | 16 | 13.91 | 44 | 38.3 |

**Table 3 (Continued)****First determination by autopsy agency** (no missingness)
|  |  |  |  |  |  |  |  |
| --- | --- | --- | --- | --- | --- | --- | --- |
| No prior deaths in custody | 242 | 49 | 20.3 | 42 | 17.4 | 89 | 36.8 |
| Prior deaths, none classified as homicide | 259 | 43 | 16.7 |  |  |  |  |
| ≥ 1 prior death classified as homicide | 439 | 176 | 40.1 |  |  |  |  |
| No prior deaths mention force-related injury | 304 |  |  | 45 | 14.8 |  |  |
| ≥1 prior deaths mention force-related injury | 394 |  |  | 68 | 17.6 |  |  |
| No prior deaths mention force or related injury | 164 |  |  |  |  | 49 | 29.9 |
| ≥1 prior deaths mention force or related injury | 534 |  |  |  |  | 262 | 49.1 |

1. Percentages refer to the percent of deaths within each stratum that are classified as homicide, mention a force-related injury/condition, or mention any force

Regarding manner of death, accidental (46.9%) was more common than homicide, while undetermined (19.5%), natural (5.0%), and suicide (0.1%) were less common. Homicide determinations increased over time, rising from 25.0% of deaths during 2012-2014 to 32.2% during 2018-2021.

For cause-of-death statements, 16.9% mentioned excited/agitated delirium and 73.9% mentioned drugs. A total of 593 deaths mentioned cocaine, amphetamines, and/or phencyclidine (PCP), comprising 89.0% of the 666 cause statements that mentioned specific drugs (29 statements referred to drugs only in general, e.g., “multiple drug toxicity”). Compared to accidental deaths, those classified as homicide were more likely to mention force (18.8% for accidental vs 74.6% for homicide) and force-related injuries/conditions (8.4% vs. 30.6%) and less likely to mention drugs (62.7% vs. 90.9%) and excited/agitated delirium (10.2% vs. 20.8%).

Concerning political affiliation, counties in the highest quartile of Republican vote share were consistently least likely to reflect force in cause and manner classifications. In unadjusted tabulations, deaths in the lowest-quartile Republican vote share counties were more likely to be classified as homicides (32.9% vs. 21.7%), mention force-related injuries/conditions (18.8% vs. 12.2%), and mention any force (48.8% vs. 30.8%) compared to the highest Republican quartile. Model-adjusted prevalence differences (lowest vs. highest Republican quartile) were 0.17 (95% CI: 0.05, 0.28) for homicide, 0.22 (95% CI: 0.06, 0.38) for mentions of force, and 0.14 (95% CI: 0.023, 0.26) for force-related injuries/conditions (Figure 1).

**Figure 1.**
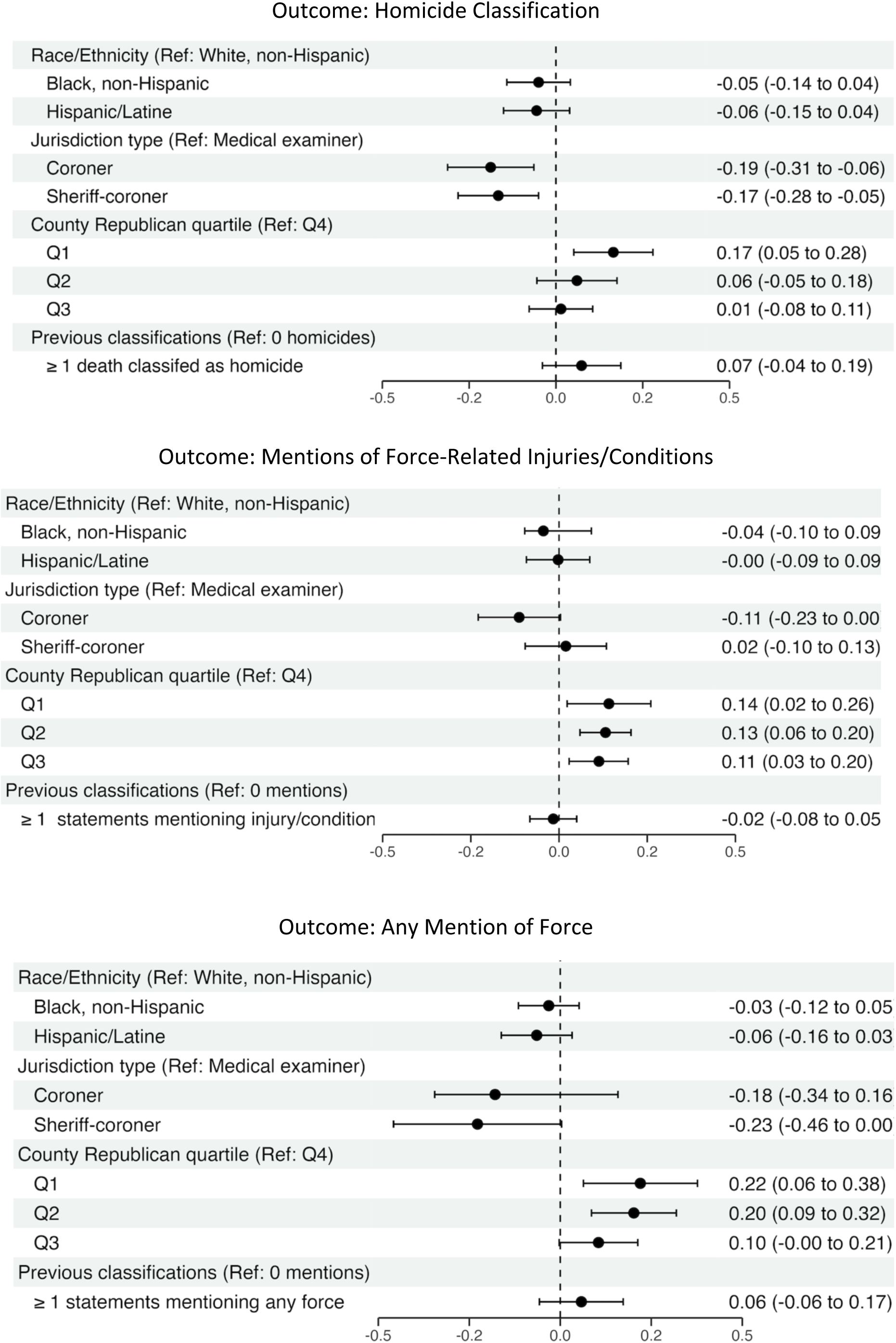
Model-Adjusted Prevalence Differences

Additionally, coroners and sheriff-coroners were less likely to classify the manner of death as homicide compared to medical examiners. The adjusted prevalence difference (reference group: medical examiner jurisdictions) was -0.19 (95% CI: -0.31, -0.06) for coroners, and -0.17 (95% CI: -0.28, -0.05) for sheriff-coroners. Differences in cause-of-death classification (force-related injuries/conditions and any mention of force) by C/ME type were inconclusive.

Regarding racial disparities, descriptive tabulations showed a notably lower proportion of non-Hispanic Black decedents were classified as homicide compared to non-Hispanic white decedents (23.9% vs. 28.4%), and non-Hispanic Black decedents were also least likely to have mentions of any force or force-related injuries/conditions. However, model-adjusted results were inconclusive across all outcomes.

Lastly, we identified idiosyncratic patterns of mortality classification that yielded inconclusive results in adjusted models. In descriptive tabulations, 40.1% of deaths were classified as homicide for agencies that had previously classified ≥1 death as a homicide vs. 16.7% for agencies that had classified all previous deaths as non-homicide. Additionally, among the 52 agencies that had ≥5 manner and cause of death classifications available, 12 agencies never assigned homicide as a manner of death, while 15 assigned homicide to more than half of deaths. For these same 52 agencies, similar evidence of agency-level variability existed for mentions of any force and mentions of force-related injuries/conditions.

## Discussion

Our national study of deaths in US police custody that follow non-firearm force found that mortality classification routinely failed to reflect the use of force that preceded the death.

Classifying deaths as non-homicide and writing cause-of-death statements that made no mention of force were widespread practices, affecting all death investigation system types, racial/ethnic groups, and local political characteristics.

Contrary to the US National Association of Medical Examiner’s suggestion to assign homicide as a manner of death for police restraint-related fatalities, fully 71.5% of the AP News Lethal Restraint incidents in our analytic dataset were categorized as non-homicide, with investigators most often choosing accidental. Death records that are neither classified as homicide nor make any mention of force comprised approximately half of the incidents in our dataset. Such non-homicide, non-force determinations may hinder efforts to identify in-custody deaths in public health databases and may make it less likely for these deaths to come to the attention of public officials, journalists, or civil society organizations.

In recent years, various US professional and legislative advocacy efforts related to medico-legal investigations of in-custody deaths have focused on banning or discouraging the use of the term excited delirium in cause of death statements.^25,26^ However, we found that excited/agitated delirium was cited relatively rarely, appearing in fewer than 1 in 5 cause-of-death statements.

Instead, it was more typical for death investigators to cite drugs or drug toxicity, which was mentioned in nearly three in four cause-of-death statements. This proportion should be considered a lower bound, as a post-mortem toxicology analysis may indicate the presence of drugs, but the pathologist may not include it as a cause of death. Since at least the 1980s, US law enforcement and medical research have identified drug intoxication, particularly when involving substances such as stimulants and PCP, as a risk factor for restraint-related deaths requiring reforms to restraint practices (e.g., avoiding neck holds and prone positioning).^27,28^ While the physiologic mechanism is unclear, recent literature has proposed a pathway by which drug intoxication increases metabolic demand and, when restraint hinders the ability to exhale sufficient, metabolic acidosis results in cardiac arrest.^29^

Our finding that coroners and sheriff-coroners were less likely to classify deaths as homicides is in line with prior research suggesting that, compared to medical examiners, sheriff-coroners are less likely to report in-custody deaths to law enforcement or public health data systems.^12^ The relative autonomy provided to medical examiners through their role as appointed professionals may, at least in part, insulate them from the political pressures that many death investigators report in surveys.^30^

The pathways by which highly Republican counties are least likely to classify in-custody deaths as homicide, mention force, or mention related conditions remains unclear. Law enforcement agency participation is essential in death investigations, and it is possible that police officials are less cooperative in counties with greater shares of Republican voters. Additionally, agencies in such counties are less likely to use body-worn cameras^31^ that may otherwise aid in investigations, and it is also possible that death investigators in these areas encounter greater political pressure to make determinations amenable to police and public officials.

Our study is subject to several limitations. First, the cause and manner of death statements were most often obtained from autopsy reports, which are produced by forensic pathologists. In many coroner jurisdictions, the coroner has authority to overrule pathologists. Individual pathologists have reported instances in which a coroner changed their homicide determination to accidental for a police-related death.^11^ It is therefore possible that we overestimate the proportion of deaths officially attributed to force in coroner jurisdictions. Second, with few deaths investigated by each agency and no data on the identity of the specific pathologist, we were unable to conduct a racial bias assessment that included within-agency or within-investigator comparisons.

The inconsistent classification of the manner and cause of deaths in police custody is an issue of public health data quality that has profound social implications. Further research may assess interventions designed to improve reporting.

## Data Availability

The data that are the basis for this study are available from https://apnews.com/projects/investigation-police-use-of-force. All data produced in the present study are available upon reasonable request to the authors

