## Supplemental File for "Mortality Classification for Deaths that Follow the Use of Non-Firearm Force by Police: A National Cross-Sectional Study (United States, 2012-2021)"

We developed a set of decision rules, which we report below, to infer the jurisdiction type for each decedent in the AP Lethal Restraint database. Our aim was to determine which agency had official legal authority over the cause and manner of death determination using available information about the incident location, state of death, and office that conducted the autopsy. We then classified the agencies with legal authority into one of three categories (medical examiner, coroner, sheriff-coroner) using an open-source data search that included, *inter alia*, reviews of agency websites, state and local death investigation statues, news reports, and, when ambiguous, direct communications with death investigation offices.

Determining the agency legally empowered to investigate the deaths was not always straightforward for several reasons. First, data provided by AP identified the agency that conducted the autopsy, but that was not always synonymous with the agency with legal jurisdiction. This is because many agencies, especially coroner’s offices, have fee-for-service arrangements for autopsies and other death investigation services (e.g., medical examiner offices in nearby jurisdictions, pro-profit companies) or the state government conducts the autopsies. We also note that, in addition to the autopsy agency variable, AP journalists made a second variable available to us, referred to as the “agency assigning manner”. However, we found that this variable was used inconsistently and did not always identify an agency that had plausible legal jurisdiction over the death investigation. Second, jurisdiction is generally based on the location of death, while the AP’s data only provided the state of death and the city incident location (i.e., the city where the interaction with police occurred). For many in-custody deaths, the individual did not die at the scene, but rather a hospital that is often located in a more urban area.

1. Jurisdiction for deaths in states with centralized medical examiner systems is always assigned to the state medical examiner’s office

For 16 states and the District of Columbia, the state’s chief medical examiner has legal authority over all death investigations. There are nuances in the organization of these state systems (e.g., whether there are regional offices and how much autonomy regional offices have), but for practical reasons we considered all these jurisdictions to be homogeneous. In addition to the District of Columbia, these states included: Alaska, Connecticut, Massachusetts, Delaware, Maryland, Maine, North Carolina, New Hampshire, New Mexico, Oklahoma, Oregon, Rhode Island, Utah, Virginia, Vermont, West Virginia.

2. Jurisdiction for deaths in states with only regional or county medical examiners is always assigned to the agency that conducted the autopsy.

In New Jersey and Florida, medical examiners operate in regions (counties and groups of counties). In Michigan and Arizona, medical examiners operate at the county level. There is no role for coroners, state medical examiners, or forensic centers in conducting autopsies in these four states. Therefore, the agency that conducted the autopsy can be treated as synonymous with the agency that had legal jurisdiction over the death investigation.

3. Jurisdiction for deaths autopsied by the state chief medical examiner’s office in Iowa, Arkansas, and Mississippi are always assigned to the state’s chief medical examiner.

In these three states, the state chief medical examiner’s office assumes jurisdiction over the death when it conducts the autopsy. In these cases, the autopsy agency and agency with legal jurisdiction are therefore synonymous.

4. Jurisdiction type for coroner-only states is categorized as coroner

These were the 9 states in which there were no either no medical examiners or the only medical examiner was a state official that played a solely advisory role: Idaho, Indiana, Kansas, Louisiana, Nebraska, Nevada, South Carolina, South Dakota, Wyoming.

5. Jurisdiction for deaths autopsied by coroners and sheriff-coroners is assigned to the autopsy agency.

We could find no evidence of coroner or sheriff-coroner agencies conducting fee-for-service autopsies for other jurisdictions. Therefore, when a coroner or sheriff-coroner conducted the autopsy, we assumed that they had legal jurisdiction over the death.

6. We individually researched the death investigation systems and deaths for decedents not covered by the decision rules above.

This included (1) deaths for which the state chief medical examiner conducted the autopsy in an advisory capacity only, and (2) states with “mixed systems”, i.e., county and regional combinations of coroner, medical examiner, and/or sheriff-coroner offices. Variables we considered in the assignment of agency jurisdiction and type of jurisdiction included the incident location, autopsy agency, AP’s “agency assigning manner” variable, news stories about the death or local death investigation system, and government websites. W left the agency name and/or jurisdiction type missing when we were not able to ascertain the variable with a high level of confidence.
